# Simplifying causal gene identification in GWAS loci

**DOI:** 10.1101/2024.07.26.24311057

**Authors:** Marijn Schipper, Jacob Ulirsch, Danielle Posthuma, Stephan Ripke, Karl Heilbron

## Abstract

Genome-wide association studies (GWAS) help to identify disease-linked genetic variants, but pinpointing the most likely causal genes in GWAS loci remains challenging. Existing GWAS gene prioritization tools are powerful but often use complex black box models trained on datasets containing unaddressed biases. Here, we use a data-driven approach to construct a truth set of causal genes in 406 GWAS loci. We train a gene prioritization tool, CALDERA, that uses a simple logistic regression model with L1 regularization and corrects for potential confounders. Using three independent benchmarking datasets of resolved GWAS loci, we compare the performance of CALDERA with three other methods (FLAMES, L2G, and cS2G). CALDERA outperforms all these methods in two out of three datasets and ranks second in the remaining dataset. We demonstrate that CALDERA prioritizes genes with expected properties, such as mutation intolerance (OR = 1.751 for pLI > 90%, P = 8.45x10^-3^). Overall, CALDERA provides a powerful solution for prioritizing potentially causal genes in GWAS loci and may help identify novel genetics-driven drug targets.

## Introduction

Retrospective analyses have found that drugs are more likely to be approved by regulators if there is human genetic evidence supporting a connection between the drug target and indication^1,2^. Indeed, 63% of drugs approved by the FDA between 2013 and 2022 were supported by human genetic evidence^3^, and the relative success of genetically-supported drug targets has not decreased over time^4^.

Genome-wide association studies (GWAS) are a valuable tool for identifying associations between diseases and genetic variants. However, the vast majority of GWAS loci contain multiple genes and the vast majority of GWAS variants do not alter protein coding sequences. A key challenge in using GWAS data to identify potential drug targets is determining which genes are affected by disease-associated variants. Several gene prioritization tools have been developed to identify the most likely effector gene for a given GWAS signal such as Ei^5^, FLAMES^6^, and L2G^7^. These three tools all model the probability that each gene in a GWAS locus is a causal gene using 1) XGBoost, 2) a truth set of causal and non-causal trait-gene pairs, and 3) a variety of features. The FLAMES study^6^ performed a head-to-head comparison of these methods and found that FLAMES outperformed L2G and Ei, which in turn outperformed cS2G^8^.

There are two main drawbacks to current gene prioritization tools. First, XGBoost models are challenging to interpret. While regression methods estimate a single effect size for each feature, the contribution of a given feature in an XGBoost model depends on the value of other variables. Second, models need to be trained on a ground truth dataset. Expert-curated causal genes have been shown to be biased toward genes in close proximity to GWAS hits and biased toward genes affected by coding credible set variants^7^. Although some methods try to mediate this by using a data-driven strategy for constructing ground-truth datasets^6^, none have actively corrected for potential sources of bias.

To address these issues, we present a novel gene prioritization tool, CALDERA (CALling Disease-RelAted genes). CALDERA uses a logistic regression model with an L1 penalty (LASSO), a small number of features, a data-driven truth set, and covariates to account for biases in this truth set. We show that CALDERA achieves state-of-the-art performance while using a simpler model.

## Methods

### Variant-to-gene evidence

We extracted predictive features for all trait-gene pairs from the original PoPS study^9^. These included distance to GWAS lead variant, non-synonymous variant PIP, ABC^10^, enhancer-promoter correlation^11–13^, eQTL colocalization^14^, PCHi-C^15,16^, SMR^17^, TWAS^18^, DEPICT^19^, NetWAS^20^, and PoPS^9^. We only included canonical ENSGIDs. To determine the number of local genes, we included all GENCODE v44^21^ genes within 300kb of the focal credible set.

### Creating a set of causal and non-causal trait gene pairs

To define a set of causal (and non-causal) trait-gene pairs, we used SuSiE credible sets for 39 independent UK Biobank GWASes^22^ (Table S1 for independent traits). To minimize the risk of errors in SuSiE fine-mapping, we subsetted to the top 5 credible sets within each region. We identified credible sets containing a non-synonymous variant with a PIP > 50% (“coding credible sets”) and the affected gene (“coding genes”). We designated the remaining credible sets as “non-coding credible sets” (no non-synonymous variant with PIP > 50%). We subsetted to non-coding credible sets within 300kb of a single coding gene for the same trait and with a maximum credible set width of 400kb. We extracted all protein-coding genes within 300kb of each of these non-coding credible sets, assigned the nearby coding gene as a “causal gene”, and assigned all others as “non-causal genes”. As such, the maximum locus size was 1Mb—a 400kb credible set plus 300kb on either side. We chose a window of 300kb because previous work has shown that 90% of eQTLs are found within 130kb of their causal gene and that 90% of GWAS hits are found within 108kb of the nearest gene (a proxy for the causal gene)^23^. We removed loci containing fewer than two genes and removed traits with fewer than five causal genes (19 traits remained). Finally, we joined variant-to-gene mapping evidence to this causal gene dataset by trait and gene.

### Feature engineering and missing data imputation

To prevent information leakage from the coding credible sets used to define causal genes, we used PoPS values that were generated using the leave-one-chromosome-out method. We left the data untransformed for PoPS^9^, coding PIPs, Andersson and Ulirsch enhancer-promoter correlations^11,13^, Jung and Javierre PCHi-C interaction scores^15,16^, DEPICT z-scores^19^, NetWAS scores^20^, and NetWAS Bon scores^20^. For TWAS^18^, we used the absolute value of the z-score. We log_10_-transformed the Roadmap enhancer-promoter correlations^12^, eQTL colocalization posterior probabilities^14^, ABC-Max scores^10^, and SMR^17^ P values. For distance-related variables (GWAS lead variant to gene body, GWAS lead variant to transcription start site [TSS]), we added 1 kilobase prior to log_10_-transformation. We used a logit_10_ transformation to convert the inverse of the number of local genes (*i.e.*, the prior probability that a gene is causal) to a log_10_ odds scale. For all log10-and logit_10_-transformed variables, we imputed missing or zero values to the minimum non-missing and non-zero value (except missing SMR P values, which were imputed to 1). For all the other variables, missing data were imputed to 0. We multiplied the transformed SMR and distance-related values by -1 to ensure a positive relationship with causal gene status.

### Relative and best-in-locus features

Within each locus, we assigned the gene with the largest value for a given feature as the “best-in-locus”, excluding ties. In addition, we constructed “relative scores” within each locus by subtracting the largest local value from each gene’s value. This resulted in a full set of 49 features: 16 groups multiplied by 3 types (global, best-in-locus, and relative), as well as the number of local genes.

### Basic feature set

We aimed to create a minimal set of features that would yield an AUPRC similar to that of the full feature set. We selected distance, coding PIP, and the number of local genes based on their importance in the L2G and FLAMES models. We selected PoPS since, unlike the aforementioned features, it integrates information from outside of the focal locus.

### Gene-level covariates

To account for bias introduced by our process for selecting causal genes, we curated a set of gene-level covariates^23^ related to genetic constraint (probability of being loss-of-function intolerant [pLI]^24^ and heterozygote selection coefficient [hs]^25^), gene length (total and coding sequence), and enhancer length (from the ABC^26^ and Roadmap^12^ datasets). We log_10_-transformed all covariate values and imputed missing values to the minimum non-missing value except pLI (missing values imputed to 0.5) and hs (missing values imputed to the maximum non-missing value). We multiplied the transformed pLI and hs values by -1 to ensure a positive relationship with causal gene status. We capped the transformed pLI at its 99th percentile (34.3) due to a long tail. We also included binary indicator variables for gene-level covariate missingness, pLI < 0.1, and pLI < 0.9.

### Model training, testing, and performance

To maximize the applicability of model predictions to new traits, we trained models using a nested leave-one-trait-out (LOTO) cross-validation framework. In the outer fold, we held one trait out as a test set. In the remaining 18 traits, we trained LASSO and XGBoost models using an inner fold of LOTO cross-validation to select hyperparameters. We used these trained models to predict causal gene probability in the held-out test set. We then used these predictions to compute AUPRC using the pr.curve function and the auc.integral method from the PPROC R package^27^. We computed AUPRC 95% CIs using the logit method^28^.

### LASSO

We trained LASSO models using the cv.glmnet function from the glmnet R package^29^, selecting the lambda value with the minimum mean cross-validated error. Where specified, we included gene-level covariates when training models but set covariate values to their mean in the held-out test sets.

### XGBoost

We trained XGBoost models using the xgboost and mlr R packages. We used a binary logistic objective function and 100 hyperparameter sets. For each set, we randomly sampled hyperparameters from uniform distributions (see Table S2 for hyperparameters and their ranges). We did not include gene-level covariates when training or testing XGBoost models.

### Recalibration

We generated calibration plots using the cal_plot_logistic function from the “probably” R package. We locally recalibrated predictions, once again using a nested LOTO framework. In each outer fold we trained models and used them to generate initial predictions in both the training set and the test set. Next, we trained a second LASSO model to predict causal gene status using the initial training set predictions (on the logit scale), as well as the relative predictions within each locus (focal -best). We applied this model to the initial test set predictions to obtain recalibrated predictions.

### Recovering known characteristics of GWAS genes

We defined putatively causal CALDERA genes as the set of 149 unique genes with a predicted causal probability > 50% for any trait. We defined putatively non-causal CALDERA genes as the remaining 2,043 unique genes in GWAS loci for these traits. Using linear or logistic regression, we tested for associations between putative causal gene status and: 1) pLI > 90%, 2) whether a gene was a transcription factor, and 3) the number of unique TSSs across gene isoforms. We extracted these gene-level features from a study by Mostafavi and colleagues^23^.

### Benchmarking

To compare the performance of CALDERA and three other tools (FLAMES, L2G, cS2G), we used three benchmarking datasets from the FLAMES study^6^: Open Targets (105 causal genes, 991 non-causal genes), ExWAS (160 causal genes, 2,019 non-causal genes), and “three molecular traits” (29 causal genes, 310 non-causal genes). We constructed CALDERA features using pre-computed values from the FLAMES study for PoPS (‘PoPS_Score’), distance (‘distance’, also used to compute the number of genes within 300kb of each GWAS signal), and coding PIP (‘VEP_sum’). Note that these coding PIP values are systematically smaller than those in the CALDERA truth set because PIP was multiplied by a shrinkage factor based on VEP effect (HIGH = 1, MODERATE = 0.6, LOW = 0.4, MODIFIER = 0.1). Seven traits in the ExWAS dataset were identical or highly correlated with the traits used to train CALDERA. We therefore used a version of CALDERA excluding these traits (calcium, estimated bone mineral density, hemoglobin, hemoglobin A1c, adult height, low density lipoprotein cholesterol, total bilirubin) for testing in the ExWAS dataset. Likewise, we used a version of CALDERA that excluded serum IGF1 protein level for testing in the “three molecular traits” dataset. We used the FLAMES, L2G, and cS2G values reported by the FLAMES study^6^.

## Results

### Defining causal genes

We constructed a set of putatively causal (and non-causal) trait-gene pairs using SuSiE^30^ credible sets for 19 independent (genetic correlation < 20%) UK Biobank traits^9^. Within a given trait, we defined causal genes as those that were 1) affected by a fine-mapped non-synonymous variant (posterior inclusion probability [PIP] > 50%) and 2) within 300kb of a separate non-coding credible set (no non-synonymous variant PIP > 50%). We defined non-causal genes as all other genes within 300kb of these non-coding credible sets. This resulted in a set of 406 putatively causal genes and 4,358 putatively non-causal genes across 19 independent traits.

### Model performance using the full feature set

Next, we trained LASSO and XGBoost models to predict causal gene status using a set of 49 features derived from: distance to GWAS lead variant, non-synonymous variant PIP (all <50% by definition), number of local genes, activity-by-contact (ABC)^10^, enhancer-promoter correlation^11–13^, eQTL colocalization^14^, promoter capture Hi-C (PCHi-C)^15,16^, summary data-based Mendelian randomization (SMR)^17^, transcriptome-wide association studies (TWAS)^18^, DEPICT^19^, NetWAS^20^, and polygenic priority score (PoPS)^9^. To assess model performance, we trained the models in a nested leave-one-trait-out cross-validation framework. Model performance in held-out traits was similar for both LASSO (Figure 1, left panel; Figure S1; area under the precision-recall curve [AUPRC] = 62.8%, 95% confidence interval [CI] = 58.0% to 67.4%) and XGBoost (AUPRC = 62.0%, 95% CI = 57.2% to 66.6). This suggests an absence of strong feature-feature interactions and non-linear relationships between causal gene status and features (after feature transformation, see Methods). Due to similar model performance, we proceeded using the simpler LASSO model.

**Figure 1.**
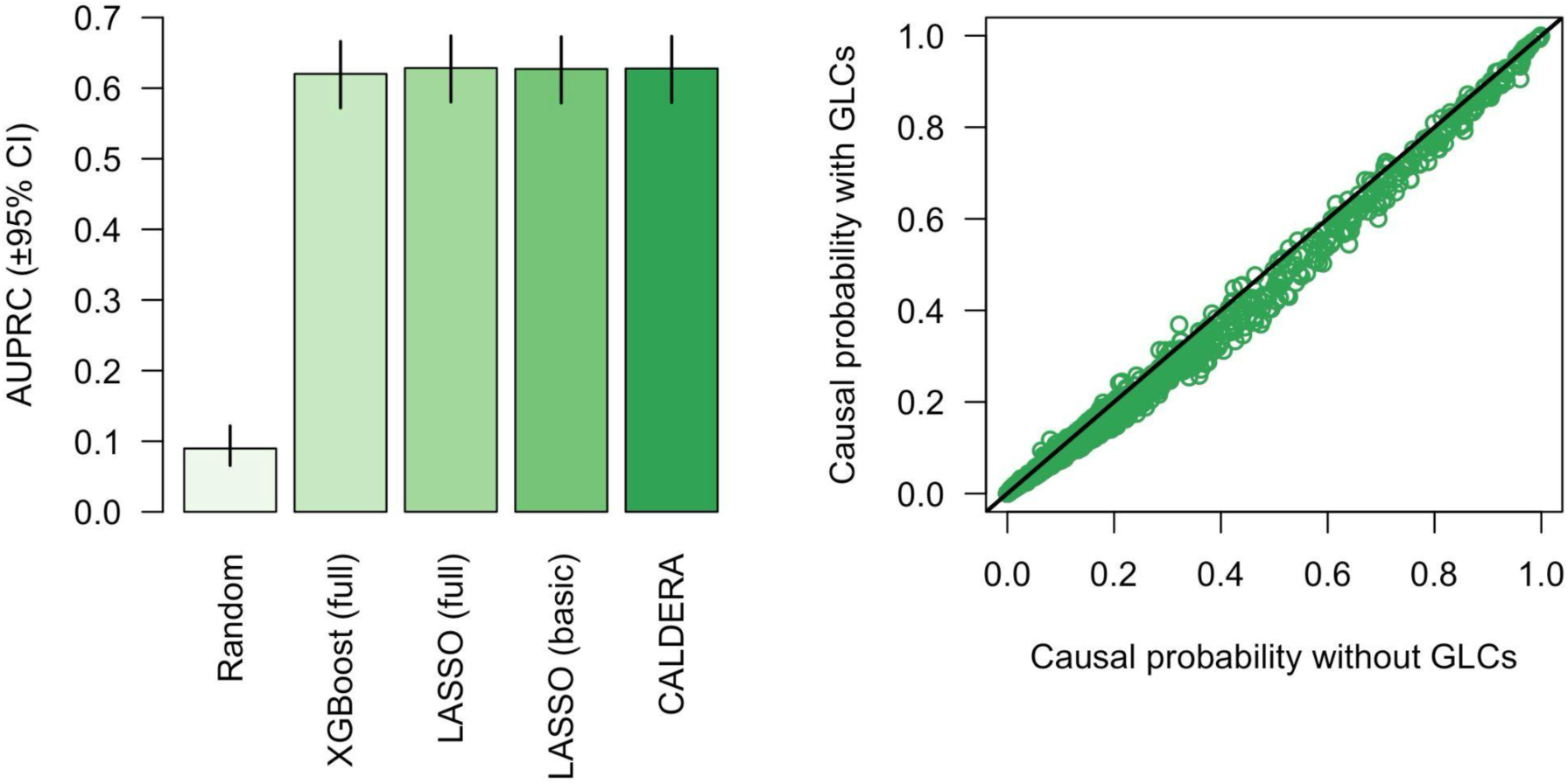
Left panel: area under the precision-recall curve (±95% confidence intervals) for models predicting causal and non-causal genes for 19 independent traits. Full = the full set of 49 gene prioritization features, basic = the basic set of 9 gene prioritization features. Right panel: causal probability estimated by LASSO models using the basic feature set with (y-axis) and without (x-axis) correcting for gene-level covariates (GLCs). Each point represents a single trait-gene pair. The solid black line represents an equivalent value for the x-and y-axis variables.

### Model performance using a basic set of features

Applying these models to obtain predictions for a new GWAS of interest requires running a wide range of pipelines to construct the full feature set. We therefore tested the performance of a LASSO model that used only a basic set of features: distance to GWAS lead variant, non-synonymous variant PIP, number of local genes, and PoPS. Despite the large reduction in the number of features, the performance in held-out traits was similar for both the full feature set (AUPRC = 62.8%, 95% CI = 58.0% to 67.4%) and the basic feature set (AUPRC = 62.7%, 95% CI = 57.9% to 67.3%; Figure 1, left panel). We therefore proceeded using the basic feature set.

### Accounting for bias

The genes nearest to GWAS lead variants (a proxy for causal genes) are more likely to be mutation-intolerant than the genes nearest to matched control variants^23^. However, we defined causal genes using fine-mapped coding variants and, therefore, our set of causal genes was enriched for mutation tolerance (Fisher’s exact test for pLI < 10%: OR = 1.725, 95% CI = 1.348 to 2.227, P = 6.0x10^-6^). A key strength of our models is the ability to account for sources of bias such as this. As such, we included a set of gene-level covariates pertaining to mutational constraint, gene length, and enhancer length. When generating predictions in the test set, covariate effects were removed by setting covariate values to their mean. Including covariates did not substantially affect model performance (Figure 1, left panel; AUPRC = 62.7%, 95% CI = 57.9% to 67.3%). After covariate correction, however, the predicted causal probabilities > 20% decreased by an average of 3.4% (Figure 1, right panel). This suggests that these predictions were inflated due to biases in the training data. We therefore performed all downstream analyses with the models trained using gene-level covariate bias correction.

### Visualizing the effects of input features

We trained a LASSO model on all 19 independent traits using the basic feature set and gene-level covariates. To help visualize the predicted feature effects, we plotted the model-predicted causal probability across a wide range of actual feature values (Figure 2).

**Figure 2.**
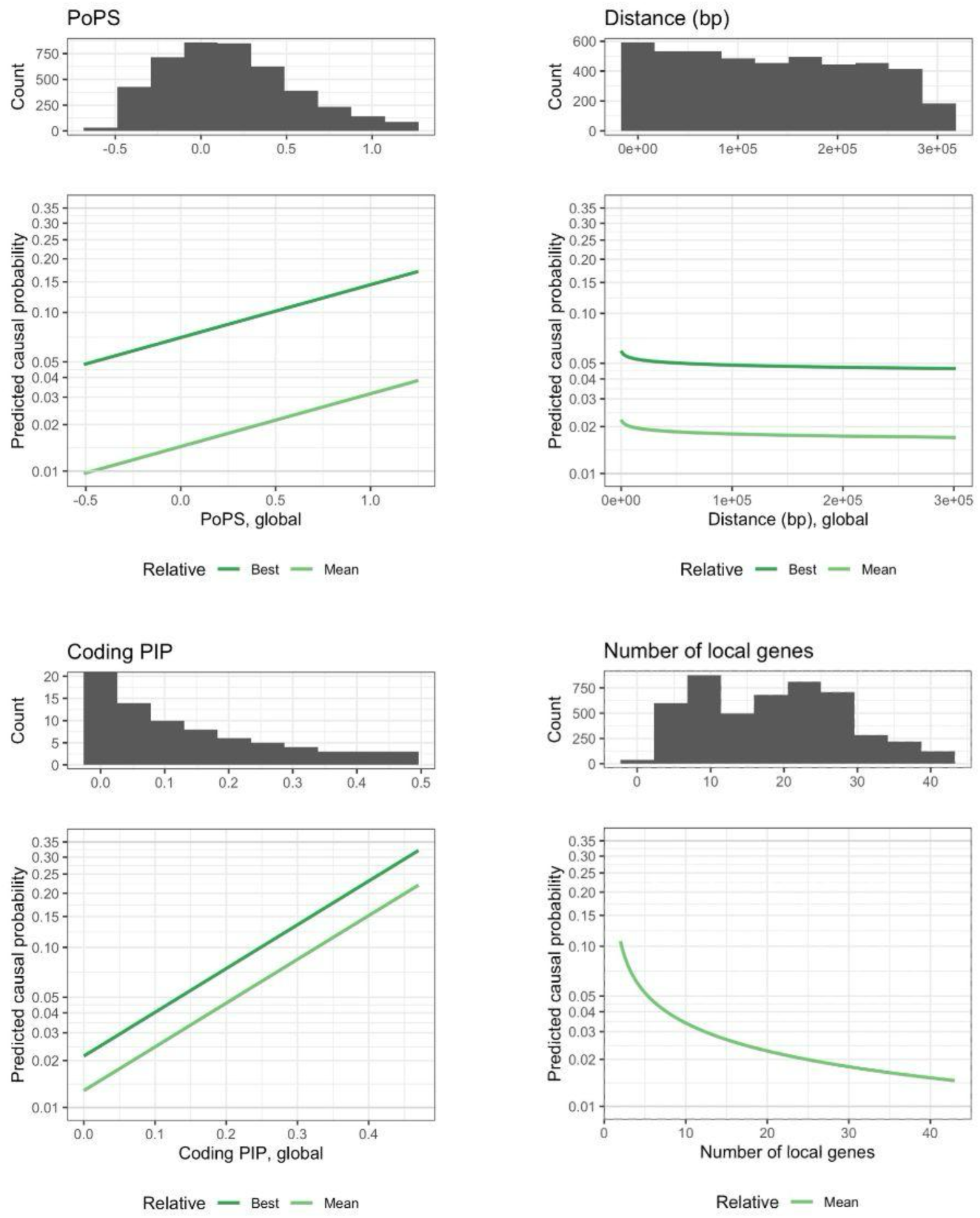
Relationships between predicted causal gene probabilities and PoPS (top left panel), distance between gene and GWAS lead variant (in base pairs, top right panel), non-synonymous credible set variant posterior inclusion probability (PIP, bottom left panel), and number of local genes (bottom right panel). The lower y-axis represents the logit-transformed probability that a given gene is causal for a given trait. The x-axis represents global feature values ranging from the 5th to the 95th percentile (except for coding PIP, which ranges from the 0th to the 100th percentile). Histograms showing the global feature distribution are plotted at the top of each panel. For coding PIP, the histogram y-axis was truncated at 20 for clarity (count of first bin = 4,787). Dark green lines represent genes with the best focal feature value in the locus. Light green lines represent genes with the average focal feature value in the locus. All other features were set to their means, leading to low overall probabilities. Although transformed distances were used to train the model, untransformed values are presented to facilitate interpretation.

### Calibration

Model predictions for held-out traits were largely well-calibrated, although predictions between approximately 25% and 55% were slightly conservative (Figure 3, left panel). Local recalibration (Figure 3, right panel; see Methods) did not negatively affect model performance (Figure 1, left panel; AUPRC = 62.8%, 95% CI = 58.0% to 67.3%) and more accurately reflected the probability that a given gene is causal for a given trait. Putting all the previous results together, we present CALDERA: a LASSO model trained on a data-driven set of causal and non-causal genes using a basic set of 9 features—as well as a set of gene-level covariates to correct for bias—followed by local recalibration.

**Figure 3.**
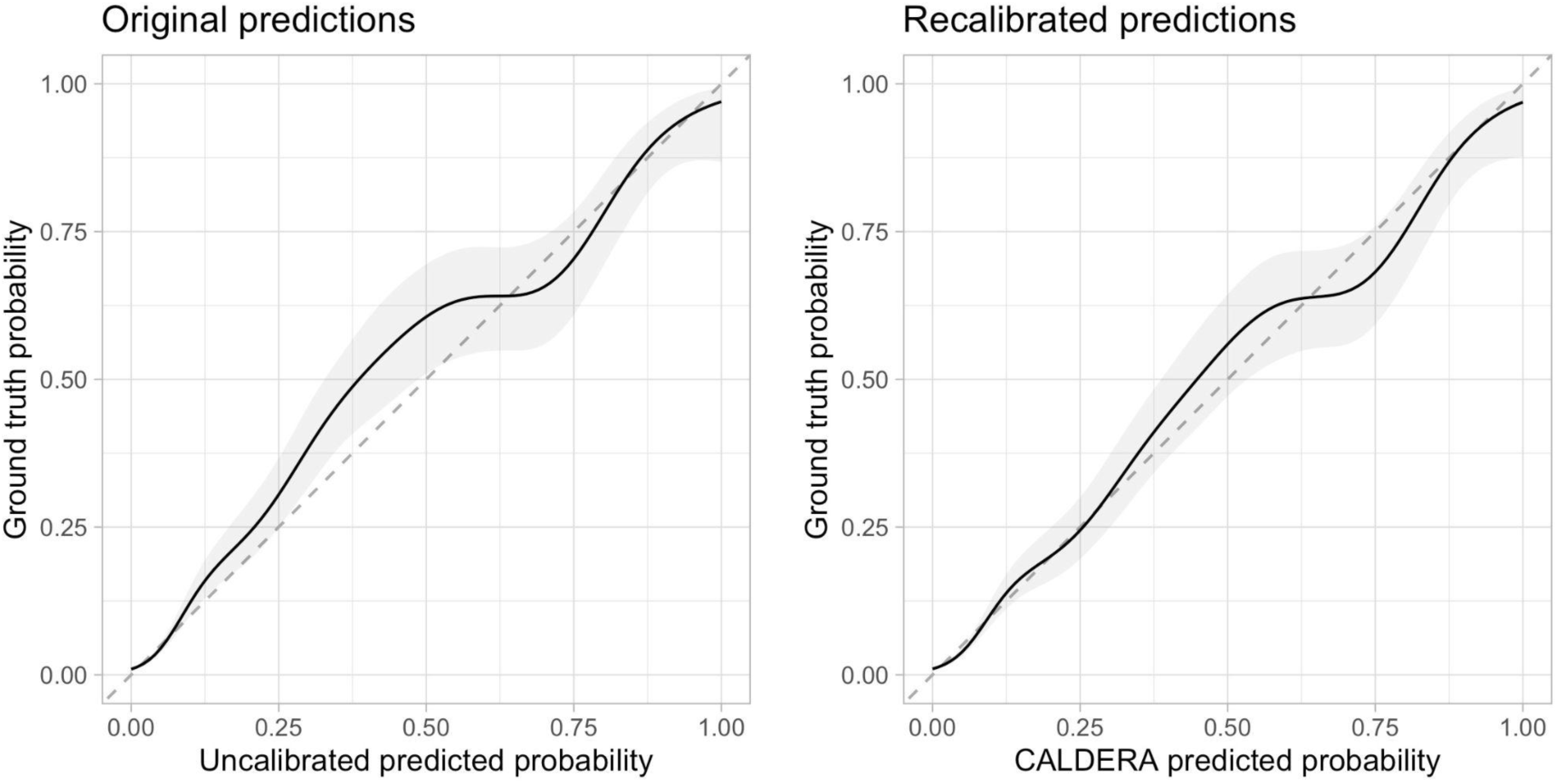
Calibration plots before (left panel) and after (right panel) recalibration. The x-axis represents the model predicted probability in held-out trait data and the y-axis represents the ground truth causal probability. The solid lines represent the fitted values from generalized additive models with shaded areas representing 95% confidence intervals. The dashed lines represent perfect calibration.

### CALDERA recovers known characteristics of GWAS genes

Previous work has shown that putative GWAS genes are more likely to be mutation-intolerant (pLI > 90%), more likely to be transcription factors, and have a greater number of unique transcription start sites (TSSs)^23^. Even though CALDERA was trained on a set of causal genes that was biased toward being mutation-tolerant, putatively causal CALDERA genes (predicted causal probability > 50% for any trait, n = 128) were more likely to be mutation-intolerant than the remaining 1,974 genes in significant GWAS loci (23.4% versus 14.7%, P = 8.72x10^-3^). We found similar results for the proportion of transcription factors (12.5% versus 6.3%, P = 8.15x10^-3^) and the average number of unique TSSs (7.3 versus 4.2, P = 2.69x10^-11^). These results demonstrate that CALDERA can effectively overcome biases in its training dataset. CALDERA prioritizes genes with expected properties and successfully recovers causal GWAS genes, even when the training set is underenriched for genes with known causal GWAS gene characteristics.

### No evidence of bias due to causal genes shared between traits

Although we only used traits with a global genetic correlation coefficient < 20%, 31 genes were causal for multiple independent traits. We repeated our analyses using a set of 189 non-shared causal genes (2,042 non-causal genes). We observed little difference in AUPRC when using models trained in the dataset without shared causal genes (Figure S3, AUPRC = 62.8%, 95% CI = 58.0% to 67.3%). Furthermore, there was a negligible difference between a logistic regression model (AUPRC = 62.8%, 95% CI = 58.0% to 67.4%) and a generalized linear mixed model using the causal gene as a random effect (AUPRC = 62.8%, 95% CI = 58.0% to 67.4%). These results suggest that CALDERA performance was not substantially inflated due to shared causal genes shared across traits.

### Benchmarking performance

We compared the performance of CALDERA with three other methods—FLAMES^6^, L2G^7^, and cS2G^8^—in three external gold standard datasets of causal and non-causal trait-gene pairs. CALDERA outperformed all other methods in the Open Targets gold standard dataset (Figure 4, left panel). Even though L2G was trained on this dataset, AUPRC was greater for CALDERA (AUPRC = 84.4%, 95% CI = 76.1% to 90.1%) than for L2G (AUPRC = 72.7%, 95% CI = 63.4% to 80.4%). In a gold standard dataset derived from burden tests of rare coding variants in the UK Biobank (Figure 4, middle panel), FLAMES outperformed all other methods (AUPRC = 55.8%, 95% CI = 48.0% to 63.3%), followed by CALDERA (AUPRC = 51.1%, 95% CI = 43.4% to 58.8%). Note that FLAMES was trained using UK Biobank rare coding variant burden data, including data from some of the same phenotypes as the test dataset. However, FLAMES was not trained on any of the loci used in the test dataset. Finally, CALDERA outperformed all other methods in a gold standard dataset derived from the serum levels of three molecules that belong to well-characterized biochemical pathways (Figure 4, right panel). CALDERA predictions were well-calibrated in all three gold standard datasets (Figure S2). These results demonstrate that CALDERA achieves state-of-the-art performance.

**Figure 4.**
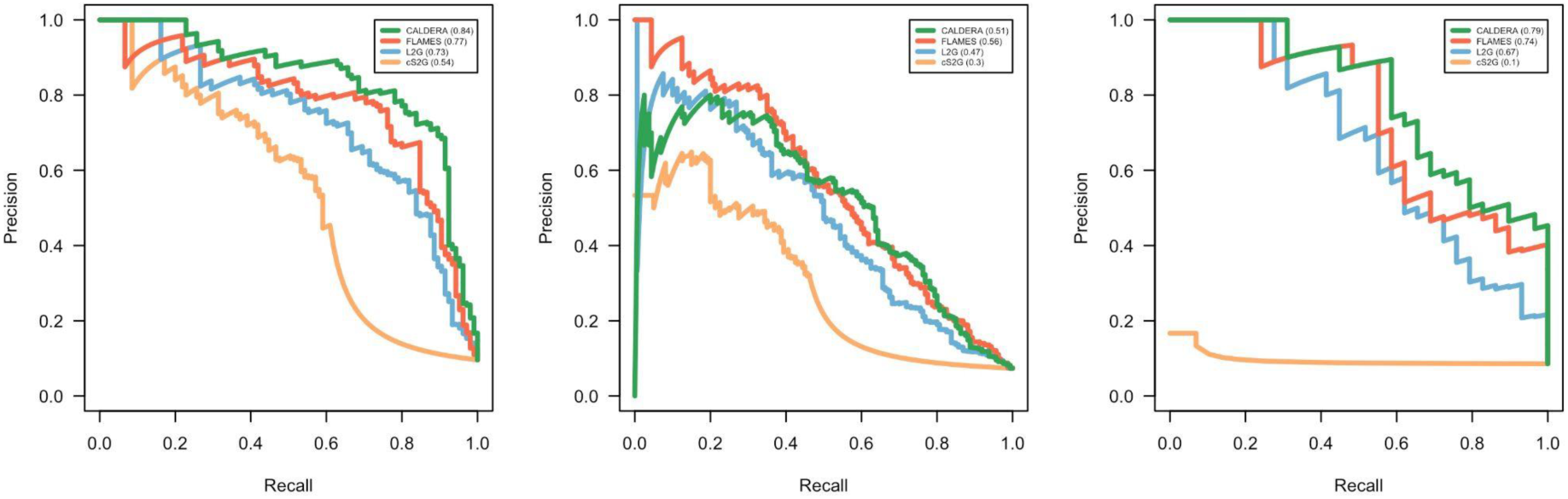
Precision-recall curves for CALDERA, FLAMES, L2G, and cS2G model predictions in the Open Targets ground truth dataset (left panel), a ground truth dataset derived from burden tests of rare coding variants in the UK Biobank (middle panel), or a ground truth dataset derived from three well-characterized serum metabolite levels (right panel).

## Discussion

In this work we have developed CALDERA, a tool for prioritizing genes in GWAS loci. CALDERA accounts for bias and achieves state-of-the-art prediction performance.

To account for biases in the CALDERA truth set, we used two strategies. First, we used a data-driven truth set rather than one that was manually curated by human experts. The L2G study found that some features related to distance and non-synonymous variants performed much better in manually-curated datasets than in data-driven datasets derived from CHEMBL^7^. This suggests that many of these causal genes were selected precisely because of their close proximity to a GWAS signal or due to a credible set coding variant. Second, we carefully considered potential sources of bias in our truth set based on how our causal genes were selected and attempted to account for these biases using gene-level covariates. To our knowledge, CALDERA is the first gene prioritization tool that attempts to actively correct for truth set biases. Failing to account for these biases led to systematic inflation of predicted causal probabilities greater than ∼20% (Figure S1). Gene prioritization tools that do not correct for biases may suffer from similarly inflated predictions. Even though the CALDERA truth set was enriched for mutation-tolerant genes, CALDERA-prioritized genes were enriched for mutation intolerance, as expected^23^.

We compared the performance of CALDERA with three other gene prioritization tools in three independent benchmarking datasets. CALDERA outperformed all other tools in the Open Targets dataset and the “three molecular traits” dataset. Notably, CALDERA outperformed L2G in the Open Targets dataset even though this was the L2G training dataset (AUPRC_CALDERA_ = 0.844, AUPRC_L2G_ = 0.727). In the ExWAS dataset, FLAMES was the only tool that achieved a greater AUPRC than CALDERA (AUPRC_FLAMES_ = 0.558, AUPRC_CALDERA_ = 0.511). In all three datasets, CALDERA and FLAMES outperformed L2G and cS2G. One possible explanation for this result is that CALDERA and FLAMES used features derived from PoPS. While most features used by gene prioritization tools are derived from a single focal GWAS locus, PoPS is a similarity-based method that integrates genome-wide information^9^. As such, PoPS may contribute information that is orthogonal to the information provided by locus-based features.

ABC, enhancer-promoter correlation, eQTL colocalization, PCHi-C, SMR, and TWAS are all locus-based features that have been shown to predict causal genes in GWAS loci^9^. However, we found that removing these tools had little impact on the predictive performance of our models. This suggests that the information provided by these methods was already accounted for by the basic set of CALDERA features derived from gene distance, non-synonymous variant PIP, PoPS, and the number of nearby genes. Removing these features allowed us to reduce the number of datasets and processing steps required to run CALDERA. We have provided code to generate CALDERA input features and output predicted causal gene probabilities using only a PoPS output file and a file containing credible set information (see Code Availability).

Since CALDERA uses a LASSO model, an increase in a given feature leads to a linear increase in the log odds that a given gene is causal. As shown in Figure 2, this approach makes it simple to visualize and understand the relationship between features and CALDERA’s predicted causal probabilities. In contrast, this is not possible for XGBoost models, where the effect of increasing a given feature is typically dependent on the values of other features. PoPS values are generated by a ridge regression model that requires ∼58,000 input features, making them challenging to interpret. Nevertheless, it is straightforward to visualize the relationship between PoPS values and CALDERA predictions.

This study has several limitations. We assumed that genes bearing a coding variant with PIP > 50% are causal for a given trait. While a variant with PIP = 50% should only have a 50% probability of being the causal variant, this probability should be much greater for coding variants^31^ and 73% of our causal genes had a coding variant PIP > 90%.

More importantly, we also assumed that all non-coding credible sets within 300kb of one of these genes also act through the same causal gene. Reprocessing published data^23^, we found that 87% of cis-eQTLs were within 100kb of their effector gene and that the percentage of effector genes decreased steeply as the distance increased further (Figure S4). We found similar results for the distance between GWAS hits and their nearest gene, a proxy for the causal gene (Figure S4). By definition, the distance between GWAS hits and their true effector genes must be greater. Nevertheless, these data and others^32^ suggest that, beyond a certain distance, the probability of being a causal gene begins to decrease in an exponential-like fashion. As such, distal causal genes in the CALDERA truth set may be less reliable than more proximal genes.

At the same time, there are well-documented examples where the causal gene lies further than 300kb from the credible set^33^. Nevertheless, CALDERA showed good calibration (Figure S2) in all three external benchmarking datasets, which used 500-750kb windows. This suggests that CALDERA can be robustly applied to larger locus definitions than the ones on which it was trained.

Another limitation of CALDERA is that it was trained on features computed using in-sample linkage disequilibrium (LD) from one cohort (UK Biobank). Using out-of-sample LD reference panels can lead to errors in both sources of CALDERA features—PoPS and fine-mapped credible sets. Additionally, GWASes that meta-analyze multiple cohorts commonly have heterogeneous sample sizes across variants. This leads to misspecified credible set PIPs^34^, although PoPS can process variant-specific sample sizes and is therefore more robust. Prior to using CALDERA, we therefore advise the use of tools to check for discrepancies between GWAS summary statistics and the LD reference panel and the removal of failing variants or loci^34,35^.

Finally, because LD patterns differ across ancestral populations, CALDERA predictions may not be well-calibrated in non-European populations. Unfortunately, this is challenging to test at present. Identifying the 406 causal trait-gene pairs in the CALDERA truth set required GWAS data for 19 independent traits, each of which was performed on hundreds of thousands of individuals. Fortunately, this is likely to be possible in the near future thanks to biobank-scale initiatives in individuals of diverse ancestries, such as All of Us^36^.

In conclusion, CALDERA is a powerful tool for GWAS gene prioritization that accounts for training set biases and uses a simple LASSO regression model. Leveraging CALDERA could aid in the prioritization of novel causal disease genes and the identification of novel drug targets.

## Data Availability

CALDERA is available as a set of open-source R scripts at https://github.com/kheilbron/caldera. All the credible set and variant-to-gene mapping data for the UK Biobank traits are available at https://www.finucanelab.org/data.
PoPS:
https://github.com/FinucaneLab/pops
MAGMA:
https://cncr.nl/research/magma/
Gencode release 44: https://www.gencodegenes.org/human/release_44.html
The Mostafavi et al. 2023 Zenodo repository: https://zenodo.org/records/6618073

https://www.finucanelab.org/data

https://github.com/kheilbron/caldera

https://github.com/FinucaneLab/pops

https://cncr.nl/research/magma/

## Acknowledgements

SR discloses support for the research of this work from the German Center for Mental Health (DZPG), the European Union’s Horizon program (101057454, “PsychSTRATA”), and The German Research Foundation (402170461, grant “TRR265”). DP and MS disclose support for the research of this work from The Netherlands Organization for Scientific Research (NWO Gravitation: BRAINSCAPES: A Roadmap from Neurogenetics to Neurobiology - Grant No. 024.004.012). DP discloses support for the research of this work from The European Research Council (Advanced Grant No ERC-2018-AdG GWAS2FUNC 834057) and the European Union’s Horizon program (964874, “REALMENT”). KH discloses support for the research of this work from the Alexander von Humboldt Foundation. DP and SR disclose support for the research of this work from the National Institute Of Mental Health of the National Institutes of Health (Award Number: R01MH124873). The content is the responsibility of the authors and does not necessarily represent the official views of the National Institutes of Health.

We thank SURF (www.surf.nl) for their support in using the Snellius National Supercomputer and Leonhard Kohleick for valuable feedback.

## Author contributions

M.S. and K.H. conceived of the study. M.S. and K.H. designed the research, performed the experiments, analyzed the data and interpreted the results. J.U., D.P, and S.R. helped advise the project. M.S. and K.H. wrote the manuscript with input from all the authors. K.H. supervised the project.

## Web resources

PoPS: https://github.com/FinucaneLab/pops

MAGMA: https://cncr.nl/research/magma/

Gencode release 44: https://www.gencodegenes.org/human/release_44.html

The Mostafavi *et al*. 2023^23^ Zenodo repository: https://zenodo.org/records/6618073

## Data and code availability statement

CALDERA is available as a set of open-source R scripts at https://github.com/kheilbron/caldera. All the credible set and variant-to-gene mapping data for the UK Biobank traits are available at https://www.finucanelab.org/data.

---

**Figure S1.**
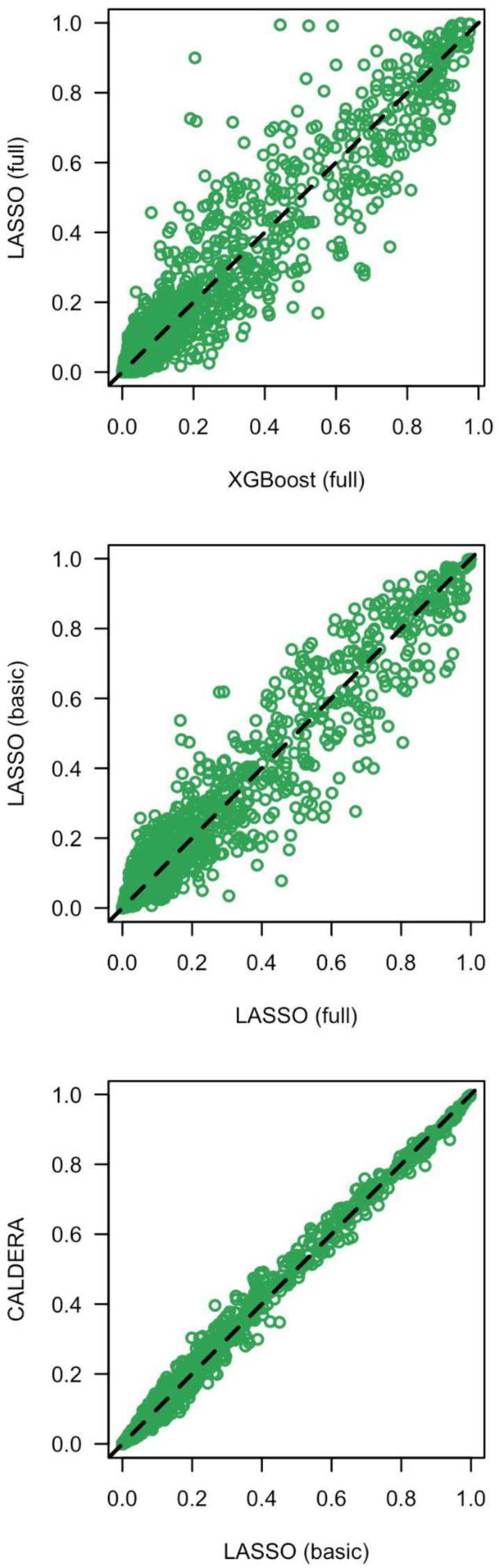
Comparisons of causal probabilities across models. **A.** XGBoost with the full feature set versus LASSO with the full feature set. **B.** LASSO with the full feature set versus LASSO with the basic feature set. **C.** LASSO with the basic feature set versus CALDERA. Each point represents a single trait-gene pair. The solid black line represents an equivalent value for the x-and y-axis variables.

**Figure S2.**
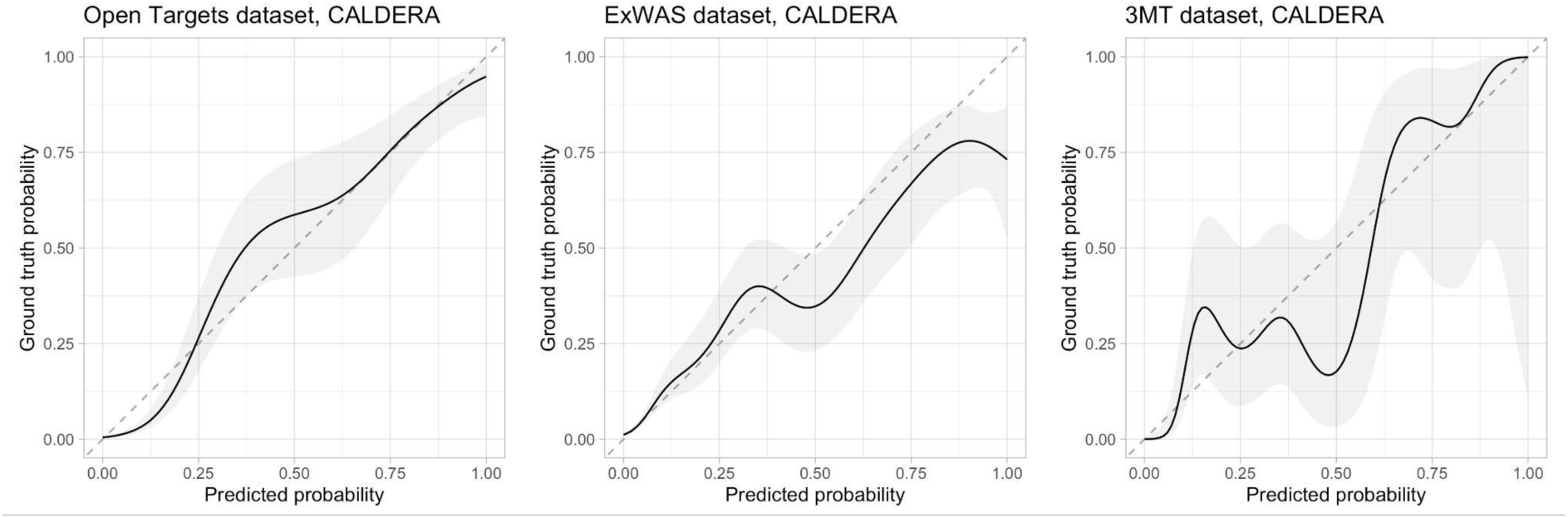
CALDERA calibration plots in the Open Targets (left), ExWAS (middle), and “three molecular traits” (right) gold standard datasets. The x-axis represents the model-predicted probability in held-out trait data and the y-axis represents the ground truth causal probability. The solid lines represent the fitted values from generalized additive models with shaded areas representing 95% confidence intervals. The dashed lines represent perfect calibration.

**Figure S3.**
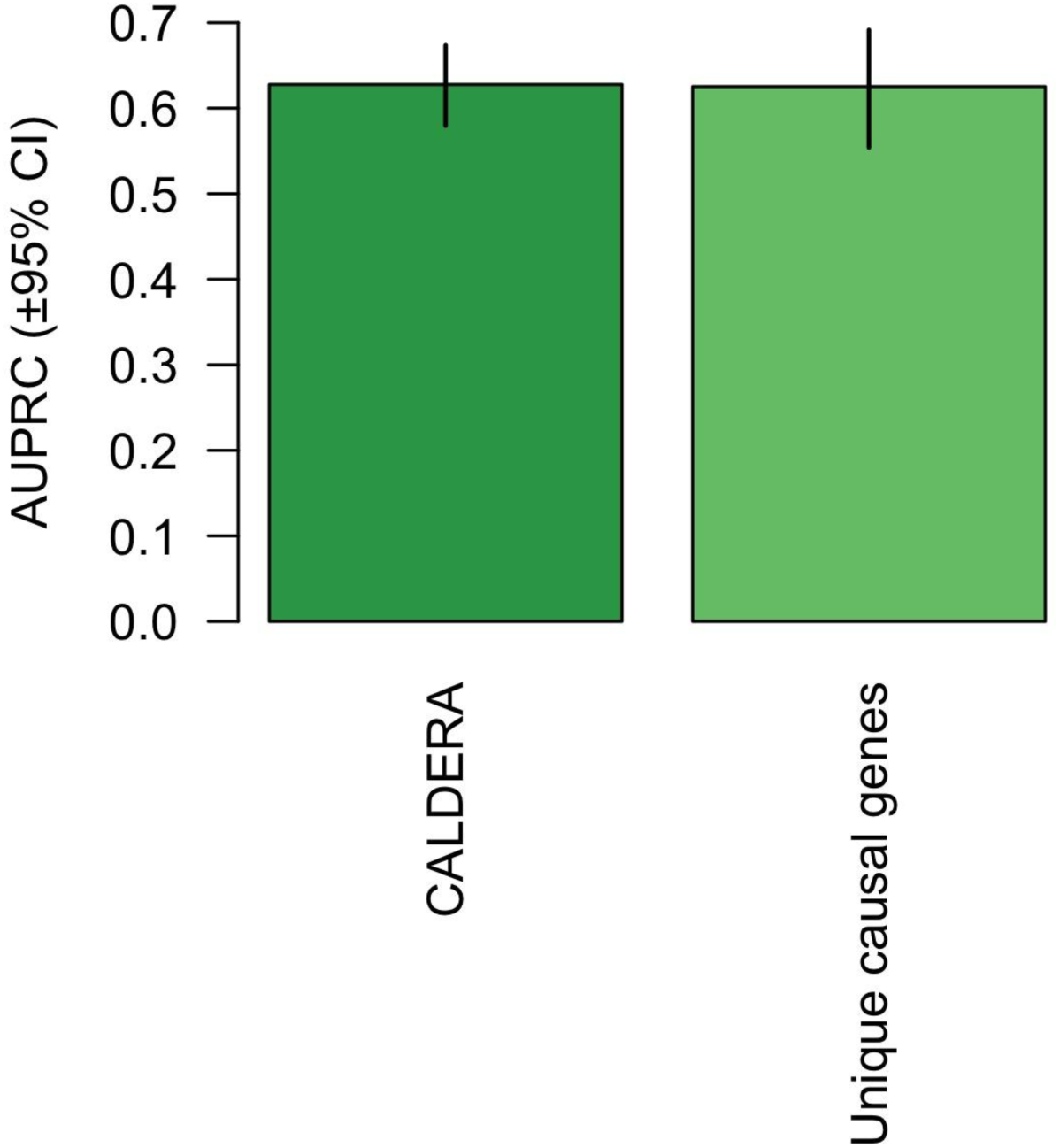
Area under the precision-recall curve (±95% confidence intervals) for models predicting causal and non-causal genes for 19 independent traits. All causal genes: results from the CALDERA model, which was trained on 406 causal genes and 4,437 non-causal genes. Unique causal genes: results from a model trained on 189 causal genes and 2,042 non-causal genes, where each causal gene is represented only once in the dataset.

**Figure S4.**
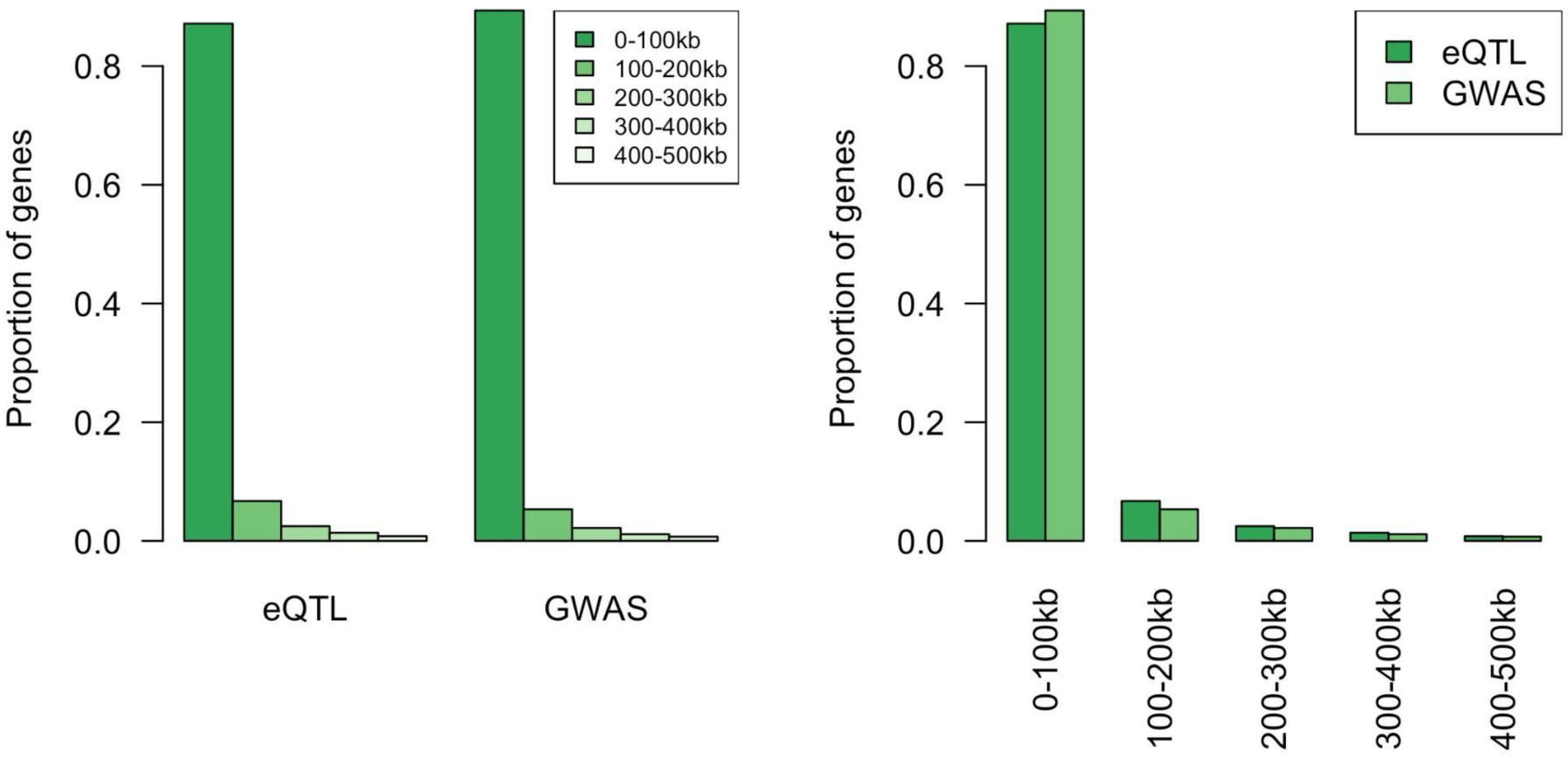
The proportion of genes that lie in various distance bins for eQTLs and their actual effector genes, and for GWAS hits and their nearest genes. The data were reprocessed from Mostafavi *et al*. 2023^23^.

